# Predictive outcome models in ischemic stroke: Comparison of Latin American Stroke Registry and RESILIENT against models

**DOI:** 10.1101/2023.03.27.23287830

**Authors:** Gabriel Torrealba-Acosta, Miguel Barboza-Elizondo, Antonio Arauz, Pablo F Amaya, Sebastian F Ameriso, Vanessa Cano, Alan Flores-Flores, Pablo M Lavados, Virginia A Pujol-Lereis, Fabiola Serrano, Sheila Martins, Raul Gomes Nogueira, Thomas A Kent, Pitchaiah Mandava

**Affiliations:** Department of Neurology, Duke University Medical Center, Durham, NC; Hospital Rafael Ángel Calderón Guardia, San José, Costa Rica; Instituto Nacional de Neurología, Mexico City, Mexico; Fundacion Valle del Lili, Cali, Colombia; FLENI, Buenos Aires, Argentina; Hospital Joan XXIII, Tarragona, Spain; Clinica Alemana De Santiago, Santiago, Chile; Universidade Federal do Rio Grande do Sul, Porto Alegre; Department of Neurology, University of Pittsburgh, Pittsburgh, PA; TAMU-HSC, Houston, TX; Stroke Outcomes Laboratory Michael E. DeBakey VA Medical Center; Baylor College of Medicine

## Abstract

**Introduction:** Despite the significant stroke burden in the region, the RESILIENT trial remains the only randomized controlled trial (RCT) for stroke treatment in Latin America. Imbalances in baseline factors typically hamper comparisons between stroke populations. The Latin American Stroke Registry (LASE) is a registry of patients receiving tPA and thrombectomy from 17 centers across 9 countries. We compared the outcomes of LASE and RESILIENT at each cohort’s baseline characteristics against models derived from other RCTs.

**Methods:** A systematic search identified RCTs that provided median NIHSS, mean age, percentage of patients receiving tPA, time-to-randomization, 90-day mRS0-2, and mortality. Akaike Information Criterion (AIC) was used to select the best model amongst 31 combinations of 5 variables. 90-day outcomes of LASE and RESILIENT were compared at their baseline values against the selected model.

**Results:** 35 RCTs encompassing 8376 subjects were identified. Models based on baseline NIHSS and the percentage of intravenous thrombolysis (IVT) were considered the most optimum in terms of AIC. The LASE registry included 950 patients receiving IVT alone, 127 that received IVT and mechanical thrombectomy (MT), and 101 receiving only MT. LASE & RESILIENT outcomes were plotted onto the models at their baseline values. LASE IVT alone group outcomes were in line with the RCT-informed model. LASE MT alone and MT + IVT, and the RESILIENT MT arm (68.5% IVT) demonstrated superior efficacy compared to no-MT. The RESILIENT control arm (71.8% tPA) had higher-than-expected mortality, while mortality of all other arms was within the predicted range.

**Conclusion:** Functional outcomes and mortality of patients from the LASE and RESILIENT trial receiving MT and MT+IVT compared favorably to the 90-day functional and mortality outcomes predicted in a model derived from no-MT data from other stroke RCTs, indicating Latin American MT systems of care are comparable to those of more resourceful regions. Higher mortality from IVT in the RESILIENT trial requires further investigation.

## 1.0 Introduction

Intravenous alteplase has been established to treat acute ischemic stroke within four and a half hours^1, 2^. Results of randomized clinical trials (RCTs) of endovascular stent-retriever (ESR) trials have shown improved functional outcomes^3-7^. Completion of ESR RCTs caused rapid changes in systems of care^8^ to adapt to this new therapy, as well as changes in guidelines from major organizations^9, 10^.

Despite a three-decade history of thrombolytic-based RCTs, questions regarding the broader applicability of treatment with alteplase to different races, ethnicities, and non-Caucasian women remain.^11^ These questions persist since the pivotal trials that led to the approval of alteplase for the treatment of stroke predominantly had Caucasian men. The thrombectomy trials^3-7^ other than RESILIENT^12^, DIRECT-MT^13^, and SKIP^14^, among others, mainly took place in North America, Europe, Australia, and New Zealand. On the other hand, the burden of stroke predominates in parts of the poorly resourced world where acute stroke RCTs have not taken place.

Prospective collection of data and maintenance of registries in different regions of the world has been offered as one solution to assessing care in areas not well represented in RCTs. However, comparison between registries and RCTs with varying baseline characteristics and modalities of treatment and selection criteria is difficult, given the importance of baseline factors in determining outcomes. Even within the RCTs, there are baseline differences between arms in factors such as baseline NIHSS^3^ and the percentage of IVT used by the various arms.^15-17^

Predictive models for outcomes, modified Rankin Score (mRS) 0-2, and mortality proportions in terms of NIHSS and age have been developed,^15, 16^ before the widespread adoption of thrombolysis. Uchino et al. also developed models for mortality in terms of NIHSS and age and NIHSS and time to treatment.^17^ These earlier predictive modeling efforts did not consider the percent of tPA use in a cohort as an independent variable^15-17^

Here we are attempting to bring information theory approaches to model development and selection of the best model in terms of loss of information from several candidate models with various combinations of variables such as NIHSS, age, percentage of IVT use, time to IV tPA, and year of publication. This approach was used to identify the best candidate model for predicting outcomes for intracerebral hemorrhage trials.^18, 19^

We aimed to compare the outcomes of a Latin American-based registry data (LASE) and RESILIENT, a thrombectomy-based trial from the same region, against a model derived from the control arms of RCTs. Such a model can be placed in the public domain so that trials with a variety of thrombectomy devices and a variable percentage of IVT can be compared against. Importantly, this approach does not require complex correction for baseline factors.^11, 15, 18, 20, 21^ A short-term (7-10 days) outcome model for stroke was previously published and placed in the public domain to serve a similar purpose. ^22^

## 2.0 Methods

### 2.1 Model development

#### 2.1.1 Literature Search

We performed a PubMed (Medline) and EMBASE search from 1994 to February 2023, using the MESH terms ‘acute’, ‘ischemic’, ‘stroke’, and ‘alteplase,’ all of them joined with the Boolean connector “AND”; followed by MESH terms ‘rt-PA’, ‘rtPA’, ‘t-PA’, joined with the Boolean connector “OR.” RCTs were chosen over non-randomized observational studies to minimize the biases of the latter type of studies. RCTs with one arm receiving intravenous tPA that reported outcomes in terms of 90-day mRS were selected. See Supplement for PRISMA flow chart.

A database of trials (see Supplement) was created that included the following information: the number of subjects in the alteplase-treated arm, median baseline NIHSS, mean age, percentage of patients that received thrombolysis, baseline glucose, time to thrombolysis (TTT) and year of publication. The database included outcomes regarding the proportion of subjects with mRS 0-1, mRS 0-2, and mRS 6 (death or mortality) at 90 days.

#### 2.1.2. Model development and selection of best models

We attempted to develop models for outcomes (mRS 0-2, mRS 6/mortality) in terms of a combination of variables found to be predictive in our prior reports, NIHSS, age, percentage of tPA use, TTT, and year of publication.^11, 15, 16, 18, 21^ With five independent variables, 31 different models are possible (i.e., 2^5^-1). The freeman-Tukey transformation was applied to the dependent variable to stabilize the variance.^15, 18, 21^ Also, to account for the wide range of subjects in the RCTs, the minimization routine was weighted by the number of subjects. Information theory approaches were adopted to select the best candidate models from the fifteen options.^18, 19^ The detailed mathematical description was provided in earlier publications field and in the Supplement as well.

Briefly, AICc, a corrected measure of information contained in the model, is calculated for each of the 15 possible combinations of models. The model with the smallest AICc is chosen as the best model.^18, 19^

### 2.2. LASE registry and RESILIENT

The LASE (Latin American Stroke Registry) is a prospectively collected dataset from January 2012 to date representing a multicenter initiative involving 17 tertiary referral centers from 9 Central and Andean Latin American countries, including Brazil, Mexico, Chile, Colombia, Costa Rica, Paraguay, Perú, Argentina, and Ecuador.^23^ All participating centers have institutional stroke programs led by stroke specialists and serve low and middle-income populations. IVT is available in all hospitals and is administered per current stroke treatment guidelines.^23^ Mechanical thrombectomy and imaging studies are not routinely available and vary from center to center. For this study, we included 1178 acute ischemic stroke subjects (950 received only IV-tPA, 101 only MT, and 127 IV-tPA and MT); all of them older than 18 and with complete-case data from the LASE registry.

The RESILIENT (Randomization of Endovascular Treatment with Stent-retriever and Thromboaspiration versus Best Medical Therapy in Acute Ischemic Stroke due to Large Vessel Occlusion Trial) was a 12-center, prospective, randomized, open-label, controlled trial conducted in Brazil. This was the first RCT enrolling only the Latin American population that compared the use of thrombectomy and IV-TPA (N=111) versus only IV-TPA (N=110) in patients older than 18 years and with a proximal large vessel occlusion.^12^

For this study, 90-day mRS 0-2 and mortality rates from both datasets were plotted onto the respective 90-day mRS and mortality predictive models. In the 3D models (Figures 1a and 1b), the middle surface defines the function and the bounding surfaces the ±90% intervals.

**Figure 1.**
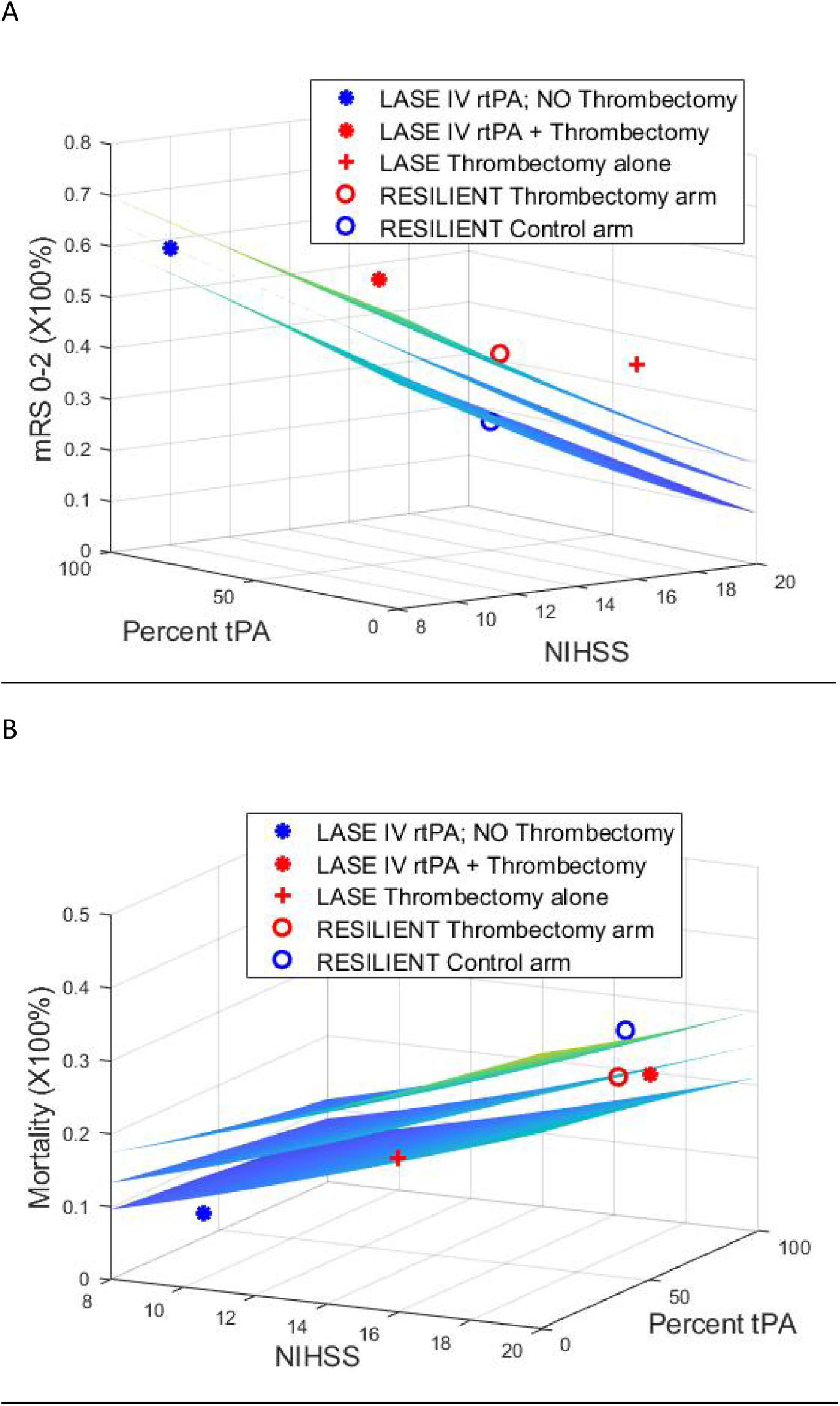
Models were generated from 35 RCT arms. The outcome (z-axis) is based on baseline NIHSS (x-axis) and percent utilization of IV tPA in an arm. Surfaces on either side represent ±p=.05 statistical intervals.). LASE IV tPA alone (blue star, N=957, 100% tPA), LASE IV tPA and thrombectomy (red star, N=127, 100% tPA), LASE thrombectomy alone (red +, N=101, 0% tPA), RESILIENT thrombectomy arm (red circle, N=111, 68.5% tPA), RESILIENT control arm (blue circle, N=110, 71.8% tPA) plotted at each trials NIHSS and percentage of tPA used. A. Good functional outcome (mRS 0-2). The LASE IV-tPA-only group was on the middle surface for mRS 0-2 (58.1%), indicating that functional outcomes aligned with the RCT-informed model. The MT only (42% mRS 0-2) and MT + tPA (46.8% mRS 0-2) and the RESILIENT MT arm (35% mRS 0-2) were above the +90% surface, indicating superior efficacy in terms of functional outcomes (mRS 0-2) when compared to IV-tPA only treated patients. b. Mortality. The LASE registry and RESILIENT arms have been plotted onto the model. LASE IV tPA-only group fell below the ±90% intervals suggesting a mortality benefit (mortality rate 8.8%). Mortality for MT only: 21%, MT+tPA: 19.5%, and RESILIENT MT arm: 24% fell within the ±90% intervals of expected mortality. Contrarily, the RESILIENT control arm had higher than expected mortality (30%), suggesting possible harm.

## 3.0 Results

### 3.1. Selection of Predictive models of stroke

We identified 35 RCT arms that met inclusion and quality criteria to develop the outcome models (See Supplement Table I). These 35 trials represented 8376 subjects with a mean NIHSS of 13.1 (range: 2-22), mean age of 68.2, and mean percent of subjects receiving IV tPA of 78.9% (range: 8.9-100%). The mean time to IVT was 146.4 minutes (range: 92-239). The median year of publication of the 35 RCTs was 2015.

#### 3.1.1 Model for modified Rankin Score 0-2

Amongst the thirty-one models considered, the model with NIHSS and percent IVT used in an arm had the lowest AICc of 211.9 (Adjusted R^2^=0.90; p<0.001; figure 1a; Supplement Table II) and was selected as the best representative model with the lowest cost of information transfer. The model with year of publication alone as an explanatory variable had a relatively low adjusted R^2^ of 0.04 (p=0.12).

#### 3.1.2 Model for mortality

Amongst the thirty-one alternate models considered, the model with NIHSS and percent tPA had the lowest AICc of 199.9 (Adjusted R^2^=0.79; p<0.001; Figure 1b, Supplement Table III) and was selected as the best representative model with the lowest cost of information transfer. The model with year of publication alone as an independent variable had an adjusted R^2^ of 0.02 (p=0.22), suggesting negligible contribution of year of publication as a predictor.

### 3.2 Plotting onto predictive models

LASE & RESILIENT outcomes were plotted onto the selected 90-day mRS and mortality models at their baseline values. LASE IV-tPA-only group included 957 subjects and was on the middle surface for mRS 0-2 (58.1%), indicating that functional outcomes aligned with the RCT-informed model. Moreover, this group fell below the ±90% intervals suggesting a mortality benefit (mortality rate 8.8%). The MT only (N=101, 0% tPA, 42% mRS 0-2) and MT + tPA (N=127, 100% tPA, 46.8% mRS 0-2), and the RESILIENT MT arm (N=111, 68.5% tPA, 35% mRS 0-2) were above the +90% surface, indicating superior efficacy in terms of functional outcomes (mRS 0-2) when compared to IV-tPA only treated patients. Mortality for these three groups (MT only: 21%, MT+tPA: 19.5%, RESILIENT MT arm: 24%) fell within the ±90% intervals of expected mortality. Contrarily, the RESILIENT control arm (N=110, 71.8% tPA) had higher than expected mortality (30%), suggesting possible harm (Fig1-B). The model predicted the mortality of the control arm of RESILIENT at a baseline NIHSS of 18 and 71.8% of IVT to be 24% (prediction intervals of 20.1-28.1%). Interactive 90-day mRS 0-2 and 90-day mortality models are provided (https://gtorrealbaacosta.shinyapps.io/90day_Models/)

## 4.0 Discussion

We successfully leveraged information theory concepts to select the best models for the functional outcome (mRS 0-2) and mortality from several alternate models derived from RCTs. Additionally, we showed that the developed models could be used to evaluate stroke outcomes of different populations by comparing them against models. These models presented here are available in the public domain, and users can interact with models to plot their study outcomes onto the model.

Since 2020, the Global Stroke Alliance and the Latin American alliance for stroke have implemented strategies to prevent and improve stroke care in LA. The number of stroke centers and access to stroke reperfusion therapies has increased during this period. Nonetheless, pre-hospital stroke care is still limited in LA countries due to a lack of organized systems; the implementation of MT is still sparse and mainly restricted to a few hospitals in each country.^24^ The RESILIENT trial demonstrated the safety, efficacy, and cost-effectiveness of implementing MT across stroke centers in the Brazilian Public Healthcare System in an under-resourced setting. Similarly, the LASE initiative strives to collect and report outcomes from stroke therapies across LA countries despite struggling to implement reperfusion therapies in countries with limited resources. In this context, it is encouraging that after comparing the functional outcomes and mortality reported in the LASE and RESILIENT onto the selected predictive models, we demonstrated the benefit of using IV-tPA and MT in patients with acute ischemic stroke in a heterogenous group of stroke centers in LA.

IV-tPA and MT efficacy may vary according to sex, race, and socioeconomic status. Correspondingly, 90-day outcomes are influenced by patient care and rehabilitation after the acute phase of stroke, which depends upon healthcare organizations and resources. Establishing the efficacy of any given intervention, as shown in our models, particularly in this context, is reassuring and informs the healthcare authorities about the returns and gains of successfully putting in place the infrastructure for access to stroke reperfusion therapies for a larger population.

The models developed before thrombolysis was widely available as a stroke treatment, and baseline NIHSS and age alone were used as predictors^15, 17^. In an earlier report considering RCT arms that used thrombolysis in all patients, we considered only NIHSS and age^16^. Here we considered additional variables of time to IVT and percent of patients that received IVT. We report that NIHSS and the percent IVT used in a trial are relatively more important factors from an information theory perspective. As a means of looking backward, the year of publication was considered, but at this time provides little predictive value. Glucose, amongst other factors considered important factors determining outcomes, is reported inconsistently, and the number of available RCTs required to build a model falls below a threshold.

### Limitations

Data from LASE are derived from the voluntary report of stroke centers and thus may lead to reporting bias, a special case of selection bias that is reportedly present in both RCTs and registry-based outcomes published in medical journals.^25^

## Data Availability

Anonymized data, as well as the statistical plan and R coding, not published within this article, will be made available by request from any investigator.

https://gtorrealbaacosta.shinyapps.io/90day_Models/

## References

1. National Institute of Neurological D, Stroke rt PASSG. Tissue plasminogen activator for acute ischemic stroke. The New England journal of medicine. 1995;333:1581–1587

2. Hacke W, Kaste M, Bluhmki E, Brozman M, Davalos A, Guidetti D, et al. Thrombolysis with alteplase 3 to 4.5 hours after acute ischemic stroke. The New England journal of medicine. 2008;359:1317–1329

3. Campbell BC, Mitchell PJ, Kleinig TJ, Dewey HM, Churilov L, Yassi N, et al. Endovascular therapy for ischemic stroke with perfusion-imaging selection. N Engl J Med. 2015;372:1009–1018

4. Goyal M, Demchuk AM, Menon BK, Eesa M, Rempel JL, Thornton J, et al. Randomized assessment of rapid endovascular treatment of ischemic stroke. The New England journal of medicine. 2015;372:1019–1030

5. Jovin TG, Chamorro A, Cobo E, de Miquel MA, Molina CA, Rovira A, et al. Thrombectomy within 8 hours after symptom onset in ischemic stroke. The New England journal of medicine. 2015;372:2296–2306

6. Saver JL, Goyal M, Bonafe A, Diener HC, Levy EI, Pereira VM, et al. Stent-retriever thrombectomy after intravenous t-pa vs. T-pa alone in stroke. N Engl J Med. 2015;372:2285–2295

7. Bracard S, Ducrocq X, Mas JL, Soudant M, Oppenheim C, Moulin T, et al. Mechanical thrombectomy after intravenous alteplase versus alteplase alone after stroke (thrace): A randomised controlled trial. Lancet Neurol. 2016;15:1138–1147

8. Suarez JI, Kent TA. The time is right to improve organization of stroke care. Neurology. 2008;70:1232–1233

9. Powers WJ, Derdeyn CP, Biller J, Coffey CS, Hoh BL, Jauch EC, et al. 2015 american heart association/american stroke association focused update of the 2013 guidelines for the early management of patients with acute ischemic stroke regarding endovascular treatment: A guideline for healthcare professionals from the american heart association/american stroke association. Stroke; a journal of cerebral circulation. 2015;46:3020–3035

10. Wahlgren N, Moreira T, Michel P, Steiner T, Jansen O, Cognard C, et al. Mechanical thrombectomy in acute ischemic stroke: Consensus statement by eso-karolinska stroke update 2014/2015, supported by eso, esmint, esnr and ean. International journal of stroke : official journal of the International Stroke Society. 2016;11:134–147

11. Mandava P, Murthy SB, Munoz M, McGuire D, Simon RP, Alexandrov AV, et al. Explicit consideration of baseline factors to assess recombinant tissue-type plasminogen activator response with respect to race and sex. Stroke. 2013;44:1525–1531

12. Martins SO, Mont’Alverne F, Rebello LC, Abud DG, Silva GS, Lima FO, et al. Thrombectomy for stroke in the public health care system of brazil. New England Journal of Medicine. 2020;382:2316–2326

13. Yang P, Zhang Y, Zhang L, Zhang Y, Treurniet KM, Chen W, et al. Endovascular thrombectomy with or without intravenous alteplase in acute stroke. New England Journal of Medicine. 2020;382:1981–1993

14. Suzuki K, Matsumaru Y, Takeuchi M, Morimoto M, Kanazawa R, Takayama Y, et al. Effect of mechanical thrombectomy without vs with intravenous thrombolysis on functional outcome among patients with acute ischemic stroke: The skip randomized clinical trial. JAMA. 2021;325:244–253

15. Mandava P, Kent TA. A method to determine stroke trial success using multidimensional pooled control functions. Stroke. 2009;40:1803–1810

16. Mandava P, Shah SD, Sarma AK, Kent TA. An outcome model for intravenous rt-pa in acute ischemic stroke. Translational stroke research. 2015;6:451–457

17. Uchino K, Billheimer D, Cramer SC. Entry criteria and baseline characteristics predict outcome in acute stroke trials. Stroke. 2001;32:909–916

18. Mandava P, Murthy SB, Shah N, Samson Y, Kimmel M, Kent TA. Pooled analysis suggests benefit of catheter-based hematoma removal for intracerebral hemorrhage. Neurology. 2019;92:e1688–e1697

19. Burnham KP, Anderson DR. Model selection and multimodel inference. Springer-Verlag New York; 2002.

20. Mandava P, Kent TA. Intra-arterial therapies for acute ischemic stroke. Neurology. 2007;68:2132–2139

21. Kent TA, Shah SD, Mandava P. Improving early clinical trial phase identification of promising therapeutics. Neurology. 2015;85:274–283

22. Torrealba-Acosta G, Barboza-Elizondo M, Fernández-Morales H, Qasim M, Litvak P, Rothlisberger T, et al. Thrombolysis experience in costa rica compared against individual patient data from two randomized controlled trials. J Stroke Cerebrovasc Dis. 2022;31:106599

23. Arauz A, Serrano F, Ameriso SF, Pujol-Lereis V, Flores A, Bayona H, et al. Sex differences among participants in the latin american stroke registry. J Am Heart Assoc. 2020;9:e013903

24. Martins SCO, Lavados P, Secchi TL, Brainin M, Ameriso S, Gongora-Rivera F, et al. Fighting against stroke in latin america: A joint effort of medical professional societies and governments. Frontiers in Neurology. 2021;12

25. Howard B, Scott JT, Blubaugh M, Roepke B, Scheckel C, Vassar M. Systematic review: Outcome reporting bias is a problem in high impact factor neurology journals. PLOS ONE. 2017;12:e0180986

